# Increased Prevalence of Rare Copy Number Variants in Treatment-Resistant Psychosis

**DOI:** 10.1101/2022.05.04.22274673

**Authors:** Martilias Farrell, Tyler E Dietterich, Matthew K Harner, Lisa M Bruno, Dawn M Filmyer, Rita A Shaughnessy, Maya L Lichtenstein, Rose Mary Xavier, Allison M Britt, Tamara F Biondi, James J Crowley, Gabriel Lázaro-Muñoz, Annika E Forsingdal, Jacob Nielsen, Michael Didriksen, Jonathan S Berg, Jia Wen, Jin Szatkiewicz, Patrick F Sullivan, Richard C Josiassen

## Abstract

**Background:** It remains unknown why ∼30% of patients with psychotic disorders fail to respond to treatment. Previous genomic investigations into treatment-resistant psychosis have been inconclusive, but some evidence suggests a possible link between rare disease-associated copy number variants (CNVs) and worse clinical outcomes in schizophrenia. Here, we test whether schizophrenia-associated CNVs are more prevalent in patients with treatment-resistant psychotic symptoms compared to previously published schizophrenia cases not selected for treatment-resistance.

**Methods:** CNVs were identified using chromosomal microarrays and exome sequencing in 509 patients with treatment-resistant psychosis (a lack of clinical response to ≥ 3 adequate antipsychotic medication trials over at least five years of psychiatric hospitalization). Prevalence of schizophrenia-associated CNVs in this sample was compared against a previous large schizophrenia cohort study.

**Results:** In total, 47 cases (9.2%) carried at least one CNV with known or possible neuropsychiatric risk. The prevalence of schizophrenia-associated CNVs (n=21; 4.1%) was significantly increased compared to a previous schizophrenia cohort study (*p* = 0.005322; OR = 1.93). This increase in prevalence was primarily due to duplications at 15q11.2-q13.1 and 16p11.2, which were independently associated with treatment-resistance in pairwise loci-based analysis.

**Conclusions:** These findings suggest that rare schizophrenia-associated CNVs, particularly duplications of 15q11.2-q13.1 and 16p11.2, may serve as biological entry points for studying treatment resistance. Further investigation will be necessary to elucidate the spectrum of phenotypic characteristics observed in adult psychiatric patients with disease-associated CNVs.

## Introduction

As many as one third of individuals diagnosed with a psychotic disorder experience treatment-resistant psychotic symptoms (TRS; commonly defined as “treatment-resistant schizophrenia”, here we consider treatment-resistance across the spectrum of psychotic disorders) ^1, 2^. These patients have chronic severe psychotic symptoms and marked impairments in social-community functioning ^3^. They experience higher rates of suicide and greater cognitive deficits than treatment-responsive patients ^4, 5^. Apart from the patients’ suffering, direct healthcare costs for TRS in the US are 3-11x higher than for the population of patients with schizophrenia as a whole, adding an estimated $32 billion annually to the national mental healthcare budget ^6^.

Currently, it is not possible to predict which individuals will respond to treatment. Numerous studies have attempted to identify clinical predictors of TRS, with limited success ^7^. Recent studies have focused on potential genetic risk factors for treatment-resistance, including the role of common risk alleles ^8-12^ and genome-wide burden of copy number variants (CNVs) ^8, 11^. These metrics contribute to the overall genetic liability for schizophrenia ^13, 14^ and may have unique contributions with regard to differences in treatment response, but so far do not provide predictive power and thus clinical utility for TRS ^11, 12^.

Apart from common variants that individually contribute minimal disease-risk, another source of genetic risk are rare, highly penetrant and/or deleterious variants ^15^. Recent studies of TRS have indicated an increased burden of rare damaging variants in mutation intolerant genes ^16, 17^ and gene sets implicated in Mendelian diseases or specific biochemical pathways, such as neurotransmission and gene targets of antipsychotic drugs ^18-20^. In CNV analysis, focusing on highly penetrant variants can involve identifying rare but recurrent CNVs impacting loci associated with clinical phenotypes (i.e., disease-associated CNVs). Disease-associated CNVs have large phenotypic effects and increase risk for a range of neurodevelopmental and psychiatric outcomes by several-fold ^21, 22^, but cumulatively effect a small percentage (∼2%) of patients with schizophrenia ^15, 21, 23^.

A recent study by Legge et al. ^10^ found no apparent excess in the prevalence of disease-associated CNVs, or of large CNVs (>500kb or >1Mb) in patients characterized as having TRS compared to general schizophrenia cohorts. However, this study, as well as others ^9, 11, 18^, used surrogate measures such as clozapine prescriptions or antipsychotic polypharmacy to define TRS without measures of clinical outcomes. While these studies make excellent use of the large deidentified datasets available to them, a major limitation is the inability to clinically confirm treatment-resistance in participants, therefore adding potential heterogeneity into the analysis. Another study ^24^, in which treatment-resistance was determined by examining clinical improvement in social/occupational functioning, found a significantly higher rate of treatment resistance (OR = 2.79) among individuals with schizophrenia carrying disease-associated CNVs.

Here, the authors sought to: 1) identify rare CNVs with known (or possible) clinical relevance in a sample of 509 patients with clinically confirmed TRS, and 2) compare the prevalence of schizophrenia-associated CNVs (SCZ CNVs) in this sample with a previously published schizophrenia cohort not selected for treatment resistance. If specific CNVs increase risk for TRS, thorough investigation may reveal clues to the biological mechanisms underlying treatment resistance. Understanding the impact of CNVs across the lifespan, including aspects of treatment-response, will also become increasingly important as a growing number of children with genetic diagnoses enter adult psychiatric services every year.

## Methods

### Subject Recruitment and Screening

Participants were recruited between April 2015 and August 2019 from five Pennsylvania state hospitals (PASH) and their affiliated long-term structured residence (LTSR) facilities. LTSRs are smaller locked facilities with 24-hour staff coverage and medication management that allow for the “deinstitutionalization” of patients back into the community but are not necessarily an indication of clinical improvement. The study was approved by the Institutional Review Boards at the Drexel University College of Medicine and the University of North Carolina at Chapel Hill, the Chief Medical Officer of the Commonwealth of Pennsylvania, Department of Public Welfare, and individual recruitment sites. Research was carried out in accordance with the Declaration of Helsinki. Written informed consent was obtained from all participants (or legal guardian with participant assent).

Subject selection was based on information derived from direct patient interviews and medical records. ***Inclusion*** criteria were: 1) ability and willingness to give informed consent (or written consent of a legal guardian with subject assent); 2) age ≥18 years; 3) current chart diagnosis of schizophrenia, schizoaffective disorder, bipolar I disorder with psychotic features, major depressive disorder with psychotic features, or psychotic disorder NOS; 4) continuous psychiatric hospitalization for ≥5 years; and 5) lack of clinical improvement despite ≥3 antipsychotic drug (APD) trials of adequate dose and duration. ***Exclusion*** criteria were: 1) a psychotic disorder associated with substance dependence; 2) a medical condition known to cause psychosis; 3) any instance of sustained treatment response. No study participants were prisoners or on forensic units at the time of enrollment. A senior psychiatrist not associated with the study (T.S., see acknowledgments) and one of the senior authors (R.C.J.) reviewed cases to confirm diagnostic inclusion/exclusion criteria (see **Supplemental Material** for additional details).

Structured interviews were conducted with subjects who were willing and able to participate (see **Table 1**). The Ammons Quick Test ^25^ was used to estimate intelligence, and the Positive and Negative Syndrome Scale (PANSS) ^26^ was administered to document current psychotic symptoms. Demographic and phenotypic data were extracted from medical records and entered into a secure electronic data capture platform (REDCap) ^27, 28^. **Figure 1** provides an overview of subject selection. From 690 initial participants, 509 were included in the final sample.

**Table 1:** Demographics of the TRS sample and CNV carriers.

| Characteristic | Entire Sample (n = 509) |  | CNV Carriers (n = 47) |  |  |  |
| --- | --- | --- | --- | --- | --- | --- |
|  | N | % | N | % |  |  |
| Sex |  |  |  |  |  |  |
| Male | 334 | 65.6 | 32 | 68.1 |  |  |
| Female | 175 | 34.4 | 15 | 31.9 |  |  |
| Ancestry |  |  |  |  |  |  |
| Hispanic | 11 | 2.2 | 1 | 2.1 |  |  |
| White | 381 | 74.9 | 42 | 89.4 |  |  |
| Black | 98 | 19.3 | 4 | 8.5 |  |  |
| Native American | 1 | 0.2 | - | - |  |  |
| Asian/Pacific Islander | 6 | 1.2 | - | - |  |  |
| Mixed Ancestry | 12 | 2.4 | - | - |  |  |
| Primary Chart DSM-IV Diagnosis <sup>A</sup> |  |  |  |  |  |  |
| Schizophrenia (all subtypes) | 238 | 46.8 | 23 | 48.9 |  |  |
| Schizophrenia, Disorganized | 14 | - | 1 | - |  |  |
| Schizophrenia, Paranoid | 119 | - | 12 | - |  |  |
| Schizophrenia, Undifferentiated | 82 | - | 6 | - |  |  |
| Schizophrenia, Unspecified | 23 | - | 4 | - |  |  |
| Psychotic Disorder NOS | 14 | 2.7 | 3 | 6.4 |  |  |
| Schizoaffective Disorder | 236 | 46.3 | 17 | 36.2 |  |  |
| Bipolar Disorder I with Psychosis | 15 | 2.9 | 3 | 6.4 |  |  |
| Major Depression with Psychosis | 6 | 1.2 | 1 | 2.1 |  |  |
| Clozapine Exposure | 259 | 50.9 | 25 | 53.2 |  |  |
| Adequate Clozapine Trial | 175 | 34.3 | 14 | 29.8 |  |  |
|  | N | Mean | SD | N | Mean | SD |
| Age (years) <sup>B</sup> | 509 | 53.6 | 13.2 | 47 | 54.5 | 13.1 |
| Age at illness onset (years) <sup>C</sup> | 479 | 21.2 | 8.7 | 45 | 22.6 | 11.7 |
| Duration of illness (years) <sup>A</sup> | 479 | 32.4 | 12.6 | 45 | 31.7 | 12.8 |
| Duration of current hospitalization (years) | 509 | 13.4 | 10.1 | 47 | 15.8 | 11.7 |
| Total number of hospitalizations <sup>A</sup> | 503 | 8.1 | 8.0 | 47 | 8.4 | 9.0 |
| Average GAF Score <sup>A, D</sup> | 472 | 31.4 | 5.3 | 46 | 30.8 | 3.4 |
| Most Current GAF | 484 | 32.3 | 6.3 | 45 | 31.1 | 5.4 |
| Modified PANSS Scores |  |  |  |  |  |  |
| Positive Symptom Score | 363 | 19.1 | 5.7 | 33 | 21.0 | 4.8 |
| Negative Symptom Score | 363 | 17.0 | 6.7 | 33 | 16.2 | 5.5 |
| Ammons Quick Test | 341 | 86.5 | 15.5 | 27 | 88.1 | 14.4 |

<sup>A</sup> Based on medical records; <sup>B</sup> age at time of blood collection; <sup>C</sup> age at onset of psychotic symptoms (hallucinations or delusions); <sup>D</sup> Global Assessment of Functioning Scale - scores range from 1 (severe impairment) to 100 (extremely high functioning).

**Figure 1:**
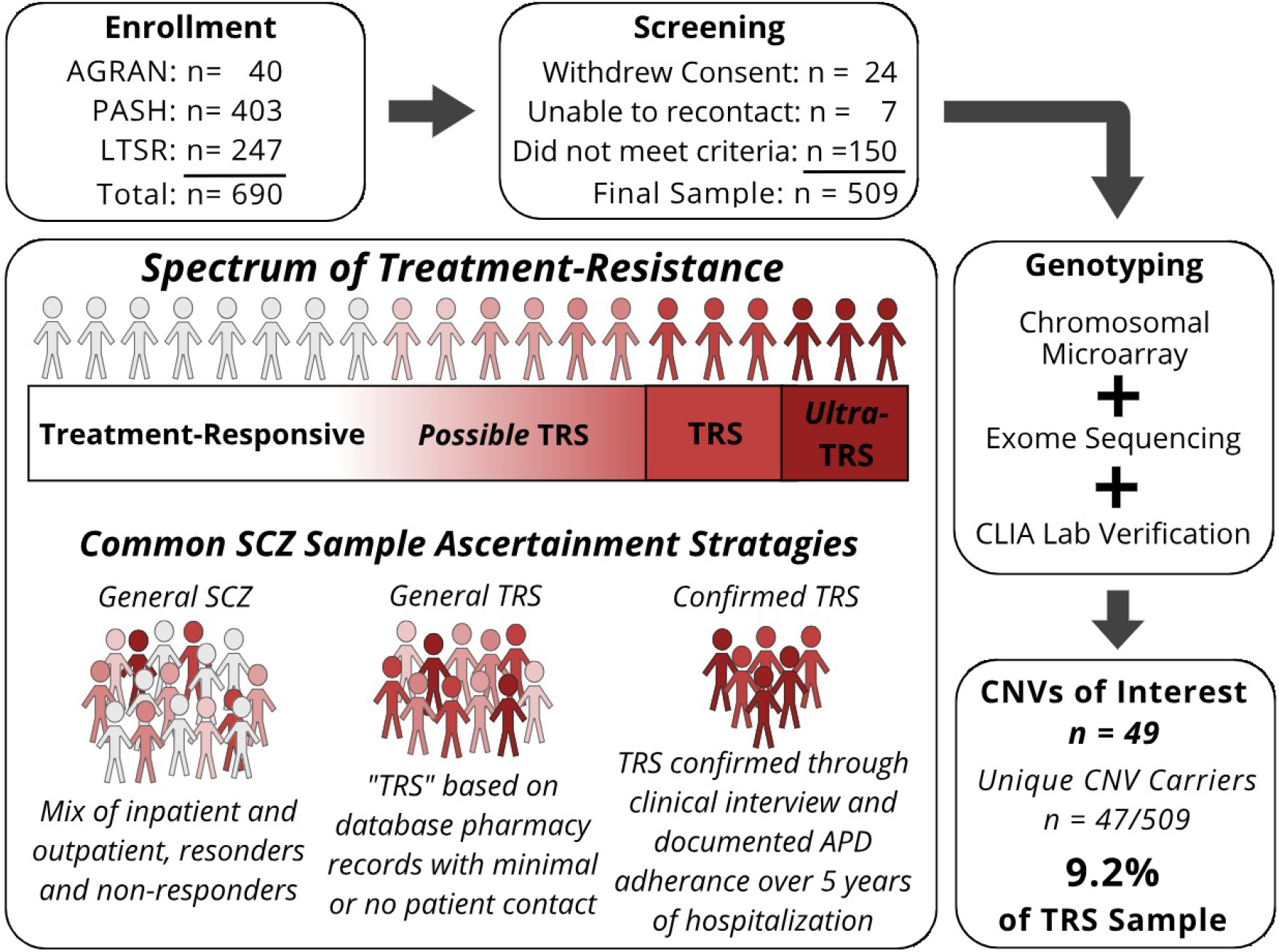
Spectrum of Treatment-Resistance and Study Workflow. Ultra-TRS describes those with TRS who also failed to respond to clozapine treatment; AGRAN, samples taken from a previous study of agranulocytosis in patients with TRS ^77^; PASH, Pennsylvania State Hospital; LTSR, long term structured residence; CLIA, clinical laboratory improvement amendment; SCZ, schizophrenia.

### Genomic Analysis

Genomic DNA was extracted from a blood sample and genome-wide SNP genotypes obtained using Illumina Infinium Global Screening Array (v2.0+MD+Psych) per standard protocols (Life & Brain GmbH, Bonn, Germany). CNVs were called from array intensity data using PennCNV ^29^, QuantiSNP ^30^, and iPattern ^31^, as packaged by EnsembleCNV ^32^. CNV calling was performed for chromosomes 1-22 and chromosome X. Individual samples were removed (n=17) due to outliers in quality control (QC) metrics (standard deviations (SD) of log R ratio (LRR), SD of B allele frequency (BAF), wave factor in LRR, BAF drift, and/or the number of CNVs detected). Additional QC steps were then followed to remove low confidence CNV calls spanning <10 probes, confidence score <10 for PennCNV, Log Bayes Factor <10 for QuantiSNP, score <1 or complex CNVs for iPattern, or with >50% reciprocal overlap with large genomic gaps (e.g., centromeres) or regions subject to rearrangement in white blood cells. We annealed adjoining CNVs that appeared to be artificially split by the CNV calling algorithm by recursively joining CNVs if the called region is ≥75% of the entire region to be joined and then removed CNVs with <100kb length. CNVs detected by two of the three calling algorithms were intersected by requiring the same subject, same CNV type, and >50% reciprocal overlap. We excluded CNVs detected by only one algorithm and merged the remaining concordant CNVs using outer probe boundaries. This approach allowed us to improve specificity (detected by ≥2 algorithms) as well as sensitivity (combining all concordant calls among three complementary algorithms). A subset of CNV calls were confirmed using a clinical-grade Agilent comparative genome hybridization array in a CLIA-certified lab (Allele Diagnostics, Spokane, WA).

Exome sequencing data were available for 478 subjects and used as a second approach to CNV analysis, and as the primary source for CNV calling in cases whose samples were excluded from SNP-based CNV analysis. Whole exome sequencing was performed using Agilent SureSelectXT Clinical Research Exome capture and was sequenced using paired end 150 bp reads on Illumina NovaSeq (GeneByGene, Houston, Texas, USA). Sequences were aligned to hg19, variants called using the GATK pipeline ^33^ and annotated using Variant Effect Predictor ^34^. CNVs were called using XHMM ^35^ and CN-Learn ^36^. For the training input to CN-Learn, previously CLIA-confirmed CNVs from the current study (n=21) along with other high confidence CNVs called from the Global Screening Array (n=10) were used (recommended minimum number of training CNVs for CN-Learn is 31). See **Supplemental Material** for diagram of CNV calling workflow.

Called CNVs were cross-referenced with a curated list of 74 CNVs associated with neurodevelopmental disorders (NDD CNVs) based on large case-control studies, including 12 associated with schizophrenia (SCZ CNVs; see **Supplemental Table S2** for complete list with genomic coordinates) ^15, 37, 38^. Large (>1 Mb) CNVs that did not overlap with known NDD CNV loci are also considered to have potential clinical value and are reported here ^39^. Three cases with CNVs >1 Mb in size were considered incidental findings and not included in this prevalence analysis as they are unlikely to be relevant to psychiatric disease. These included a mosaic copy loss of the Y chromosome, a 17p12 deletion associated with Hereditary Neuropathy with Liability to Pressure Palsies, and a 16p11.2-p11.1 duplication in a region non-sensitive to copy number dosage changes (hg19 chr16:34,197,143-35,257,261; https://dosage.clinicalgenome.org/).

### Statistical Analysis

To test whether SCZ CNVs, in aggregate, increase risk for TRS, we compared the prevalence of SCZ CNVs in this sample to previously published data from a large schizophrenia sample (21,094 subjects not selected for treatment resistance) ^15^ using a chi-squared test of independence. These same data were used to test for associations with individual SCZ CNVs using 2-sided Fisher exact tests, with p-values adjusted to correct for multiple testing using Benjamini–Hochberg false discovery rate (BHFDR; adjusted p-values <0.05 were considered statistically significant). Only variants observed at least once in this TRS sample, and for which data were available in Marshall et al. ^15^ (CNV probe-level results), were included in loci-based analysis. Additionally, we checked for demographic differences between CNV carriers and non-carriers across all variables listed in **Table 1**; only Positive PANSS scores were significantly different between groups *t*(361) = 1.9958; uncorrected p = 0.0467), with CNV carriers (*M* = 20.97; *SD =* 4.824967) having higher average scores than non-carriers (*M* = 18.91; *SD =* 5.72927). All statistical analyses were performed using R Statistical Software (v3.6.3) ^40^ and the epitools R package (v0.5-10.1) ^41^.

## Results

Demographic data are displayed in **Table 1**. Of 509 participants, 47 (9.2%) carried at least one CNV with potential relevance to their clinical presentation, which were grouped as follows: 1) 24 patients (4.7% of the sample) carried CNVs previously associated with neurodevelopmental and/or psychotic disorders; 2) 11 patients carried large CNVs (>1 Mb) which did not overlap with NDD/SCZ CNV loci, but nonetheless may impact their clinical presentation due to their size ^39^; and 3) 12 cases carried variants of uncertain significance (VUS), including CNVs partially overlapping (<50% coverage) with known NDD CNV loci and duplications of regions where only deletions are known to increase disease risk.

### Known Risk CNVs for Neurodevelopmental/Psychotic Phenotypes

Of 24 NDD CNVs identified, 21 affected one of the 12 SCZ CNV loci (see **Table 2**). The prevalence of SCZ CNVs in this TRS sample (4.1%) represents a statistically significant increase compared to a large previously published schizophrenia cohort (2.2%) ^15^, (χ2 (1, N = 21,603) = 7.77, p = 0.005322; OR = 1.93; 95% CI, 1.24-3.02). Importantly, the combined prevalence of these 12 SCZ CNVs has been replicated with relatively consistent findings across multiple schizophrenia samples (∼2.1 - 2.3%) ^21, 24, 37^. This is indicative of an association between this group of CNVs and increased risk for treatment resistance in psychosis.

**Table 2:** CNVs Associated with Neurodevelopmental/Psychotic Phenotypes.

| SCZ CNVs | Canonical CNV coordinates<br>(size in kb) | Cases<br>(N) | Case CNV coordinates<br>(size in kb) | Source | Prevalence (%) |  | P Value |  |
| --- | --- | --- | --- | --- | --- | --- | --- | --- |
|  |  |  |  |  | TRS<br>(n=509) | SCZ <sup>15</sup><br>(n=21094) | Uncorrected | Adjusted<br>for FDR |
| 3q29 Del | chr3:195.72-197.35 (1,635) | 1 | chr3:195.8-197.1 (1,302) | CMA, CLIA | 0.20 | 0.076 | 0.333 | 0.389 |
| 7q11.23 Dup | chr7:72.74-74.14 (1,398) | 1 | chr7:72.74-74.14 (1395) | CMA | 0.20 | 0.062 | 0.284 | 0.389 |
| 15q11.2 Del | chr15:22.81-23.09 (298) | 3 | chr15:22.82-23.09 (263) | CMA, CLIA | 0.59 | 0.45 | 0.504 | 0.504 |
| 15q11.2-13.1 Dup | chr15:22.81-28.39 (5,585) | 4 | chr15:23.72-28.51 (4,796) | CMA, CLIA | 0.78 | 0.071 | 0.000891 | 0.00624 * |
| 16p11.2 (distal) Del | chr16:28.82-29.05 (224) | 1 | chr16:28.49-29.05 (558) | CMA, CLIA | 0.20 | 0.052 | 0.249 | 0.389 |
| 16p11.2 (proximal) Dup | chr16:29.65-30.20 (550) | 6 | chr16:29.66-30.19 (535) | CMA, CLIA, WES | 1.18 | 0.30 | 0.00569 | 0.0199 * |
| 16p13.11 Dup | chr16:15.51-16.29 (782) | 1 | chr16:15.12-16.29 (1,162) | CMA, CLIA | 0.20 | 0.40 <sup>A</sup> |  |  |
| 22q11.21 Del | chr22:19.04-21.47 (2,429) | 4 | chr22:18.92-21.46 (2,541) | CMA, CLIA | 0.78 | 0.28 | 0.0583 | 0.136 |
| NDD CNVs |  |  |  |  |  |  |  |  |
| 1q21.2 Dup | chr1:145.39-145.81 (413) | 1 | chr1:145.4-145.8 (412) | WES |  |  |  |  |
| 15q13.3 Dup | chr15:30.94-32.52 (1,572) | 1 | chr15:31.16-32.44 (1,280) | CMA, CLIA |  |  |  |  |
| 16p12.2 Del | chr16:21.95-22.43 (482) | 1 | chr16:21.95-22.43 (478) | CMA, CLIA |  |  |  |  |
| Total case count (% of TRS sample, n = 509) |  | 24 (4.7%) |  |  | 4.13 | 1.95 <sup>B</sup> |  |  |

To identify associations of individual SCZ CNVs with TRS, we independently tested the prevalence of each SCZ CNV in our sample against that observed in Marshall et al. ^15^. Duplications at 15q11.2-q13.1 and 16p11.2 were significantly associated with TRS after correction for multiple testing. We found a greater than 11-fold increase in the prevalence of 15q11.2-q13.1 duplications (0.78%; corrected p = 0.00624; OR = 11.1; 95% CI, 3.68-33.66) and an almost four-fold increase in the prevalence of 16p11.2 duplications (1.18%; corrected p = 0.0199; OR = 3.98; 95% CI, 1.72-9.24). When these two CNVs were removed from the aggregate analysis, there was no longer a statistically significant increase in the combined prevalence of SCZ CNVs (χ2 (1, N= 21,515) = 0.2195, p = 0.6394), suggesting that these CNVs drive the collective increase in prevalence of SCZ CNVs in this sample.

### Additional CNV Findings

As shown in **Table 3**, 11 participants (2.2% of the total sample) carried large CNVs (>1 Mb) in regions not previously associated with neurodevelopmental/psychiatric phenotypes in case-control studies (one case carried two large CNVs). For most of these loci, case reports exist which describe neurodevelopmental phenotypes in individuals with overlapping CNVs, including one CNV which was previously reported in patients with psychotic symptoms (see **Supplemental Table S3**). Many of these loci involve genes highly expressed in the brain or have been implicated in central nervous system or neurodevelopmental disease (**Table 3**).

**Table 3.**
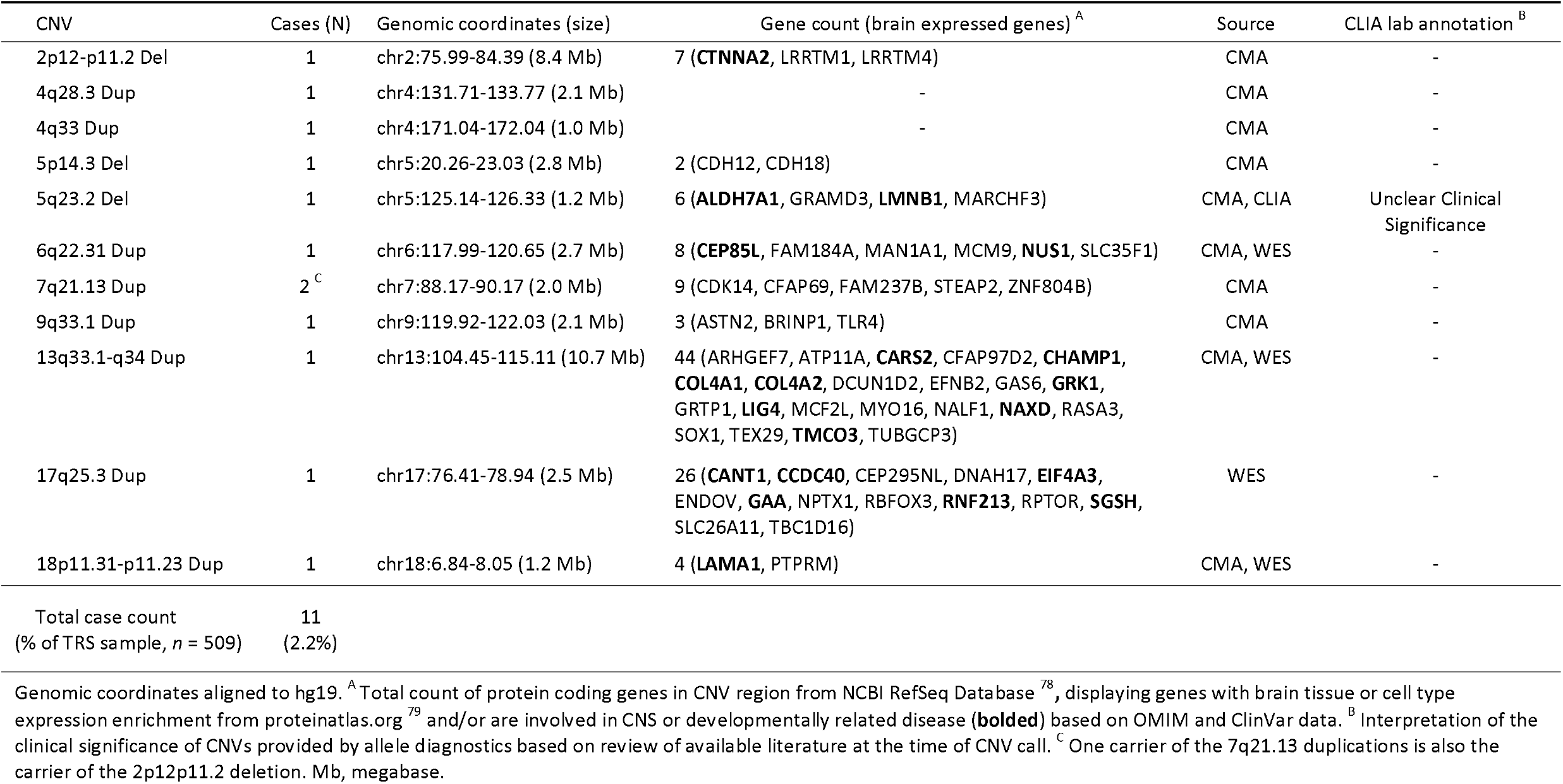
Large (>1Mb) CNVs not implicated in neurodevelopmental phenotypes in case-control studies.

Finally, 12 participants (2.4%) carried one or more VUS CNVs which do not fit the above categories but nevertheless may influence phenotypic expression in carriers (see **Table 4**). Seven carried duplications in regions where deletions are known to be associated with neurodevelopmental phenotypes, including 2q37, 15q11.2, 16p12.2, and distal 22q11.2. Six carried CNVs which overlapped with known NDD risk CNVs by less than 50%, including deletions of 1q21.1 and 22q11.2, as well as deletions and duplications of 15q13.3. These regions have known susceptibility to structural rearrangements that vary in size and breakpoint location due to the presence of several genomic low copy repeats, with some data supporting the phenotypic effects of smaller or atypical CNVs ^42-45^. One case was a carrier of both 15q13.3 and 2q37 duplications.

**Table 4:** Variants of Uncertain Significance with Potential Relevance to Clinical Phenotypes.

| CNV | Cases (N) | Genomic coordinates (size) | Source | CLIA lab annotation <sup>A</sup> | CNV group <sup>B</sup> |
| --- | --- | --- | --- | --- | --- |
| 15q11.2 Dup | 4 | chr15:22.78-23.23 (449 kb) | CMA, WES | - | 1 |
| 16p12.2 Dup | 1 | chr16:21.96-22.59 (623 kb) | CMA, WES | - | 1 |
| 2q37 (HDAC4) Two Copy Gain | 1 | chr2:239.98-240.26 (270 kb) | CMA, CLIA | Unclear Clinical Significance | 1 |
| 15q13.3 (CHRNA7) Del | 1 | chr15:32.03-32.44 (409 kb) | CMA, CLIA | Pathogenic | 1 |
| 15q13.3 (CHRNA7) Dup | 3 <sup>C</sup> | chr15:32.03-32.44 (409 kb) | CMA, CLIA | Population Variant | 1 |
| 1q21.1 (proximal TAR region) Del | 1 | chr1:145.64-145.80 (163 kb) | CMA | - | 2 |
| 22q11.2 (distal) Dup | 1 | chr22:22.13-22.58 (454 kb) | CMA | - | 2 |
| 22q11.2 Del | 1 | chr22:21.06-21.41 (358 kb) | CMA, WES | - | 2 |
| Total case count<br>(% of TRS sample, *n* = 509) | 12<br>(2.4%) |  |  |  |  |

## Discussion

Treatment-resistant psychosis is a complex and severe form of neuropsychiatric disease for which novel therapeutic insights are expeditiously sought after. Several clinical variables (i.e. age of onset, social dysfunction) may have potential for predicting treatment response ^10^, but do not directly lend themselves to novel mechanistic insights into TRS. Here, we identified an increased prevalence of schizophrenia-associated CNVs in our TRS sample compared to schizophrenia patients not selected for treatment-resistance. This is consistent with findings from a previous schizophrenia cohort ^24^ that indicated an increased rate of treatment-resistance in patients carrying disease-associated CNVs versus non-carriers.

An important consideration is whether the increased prevalence of SCZ CNVs identified in this sample is: 1) the result of specific CNVs affecting downstream biological pathways with direct relevance to treatment resistance etiopathology (i.e., dopamine system functioning, APD pharmacology, or other neurotransmitter systems such as glutamate or GABA signaling) ^46^; or alternatively, 2) a secondary result of more generalized effects of CNVs resulting in reduced social or cognitive functioning that indirectly impact treatment outcomes. This question is akin to separate models (summarized by Mulle et al.) ^47^ that view CNVs as having either specific risk for specific diagnostic outcomes, or generalized risk for neuropsychiatric disorders more broadly.

### Generalized vs Specific Risk

From the perspective of generalized risk, any effect that disease-associated CNVs exert on TRS risk may be mediated first through early neurocognitive impact. There is compelling evidence for neurodevelopmental factors being implicated in TRS. Treatment resistance has been genetically correlated with traits related to intelligence and cognition in terms of having overlap in polygenic risk alleles ^12^. Studies have also found lower premorbid IQ, younger age at onset, and poorer premorbid social adjustment are associated with TRS ^10, 11, 48^. Similar clinical features are also predictive of which individuals with schizophrenia carry clinically relevant CNVs. Specifically, a high prevalence of disease-associated CNVs are found in schizophrenia patients with comorbid intellectual disability (24%; IQ < 70), and/or childhood onset psychosis (11.9%; onset < 13 years of age) ^11, 49, 50^.

A report from Kushima et al. ^24^ provides more direct evidence that neurodevelopmental burden in CNV carriers may be related to APD response. Among patients with schizophrenia carrying disease-associated CNVs, treatment resistance was seen more than twice as often in patients with co-occurring congenital and/or developmental phenotypes (48% had TRS) versus those without developmental phenotypes (21.4% had TRS). Importantly, no CNVs have been identified which increase the risk for psychosis alone. All SCZ CNVs also increase risk for various other neurodevelopmental disorders (e.g., developmental delay, intellectual disability, autism spectrum disorder), and often appear at higher rates in neurodevelopmental cohorts than in schizophrenia cohorts. In combination with the findings from our TRS sample, these data suggest that SCZ CNVs may offer unique opportunities to study processes and shared mechanisms between neurodevelopmental burden, psychosis, and APD response.

Conversely, there is an important aspect of specificity in the contribution of CNVs to neuropsychiatric outcomes, especially regarding psychotic phenotypes. Of the most highly established NDD CNVs, only a small subset confer risk for schizophrenia ^15, 37^. It is likely that the distinct effects that CNVs have on neurobiology underly such genotype-specific outcomes ^51, 52^. From the 74 NDD CNVs that we screened for (**Table S2**), 88% (21/24) of the NDD CNVs identified in our TRS sample affected one of the 12 SCZ CNV loci (**Table 2**). Furthermore, the overall increase in SCZ CNVs observed in this sample is driven by duplications at only two loci; 16p11.2 and 15q11.2-q13.1. Also of note, CNVs occurring within the larger 16p13.11-p11.2 and 15q11.2-q13.3 genomic regions collectively affect 5.1% of our sample and account for 26 of the 47 CNV carriers in our sample overall (n = 10 and n = 16 with 16p13.11-p11.2 and 15q11.2-q13.3 CNVs, respectively). If replicated, these two CNV loci may represent important sites to focus exploration into TRS biomechanisms.

### 15q11.2-q13.1 and 16p11.2 Duplications

Duplications at 16p11.2 and 15q11.2-q13.1 are both associated with variable early clinical outcomes such as global developmental delays, neurological disturbances, and autism spectrum disorder among others) which have been extensively studied ^53-59^. Beside a few isolated reports, however, the longitudinal effects of these two variants have remained largely unexplored in clinically affected adults, particularly regarding APD response. Kushima et al. ^24^ identified 16p11.2 duplications in three adult patients with schizophrenia, of which two were reported as treatment-resistant with prominent positive symptoms, while the third (age 18) had no history of antipsychotic treatment. Another 16p11.2 duplication carrier (age 18) reported by Filges et al. ^60^, presented with psychiatric symptoms including anxiety, mood lability, and paranoid ideation which were minimally responsive to treatment with quetiapine and sertraline. Our findings add six additional 16p11.2 duplication cases with TRS to the literature.

To our knowledge, no reports exist which document APD treatment response in adult 15q11.2-q13.1 duplications carriers, although one study ^61^ indicated a relative inefficacy of benzodiazepines in the treatment of seizures in 15q11.2-q13.1 duplication carriers, which was hypothesized to be related to disruption in GABAergic signaling (several GABAβ3 receptor genes are located in the 15q11.2-13.1 region). Likewise, mouse models of the 16p11.2 duplication highlight behaviorally-relevant changes in to GABAergic interneuron and cortical pyramidal neuron activity leading to excitatory-inhibitory imbalances ^62-64^. Disruption in GABAergic inhibitory signaling has previously been implicated in psychosis and APD response, and thus may represent one possible convergent mechanism of these CNV sites for TRS ^65, 66^. Importantly, both regions contain several dozen genes, and therefore mechanisms of action related to APD response may be complex and multigenic. Yet, our findings suggest that 16p11.2 and 15q11.2-q13.1 duplications may provide opportunities to model treatment resistance within experimental systems, with the aim of uncovering bio-mechanisms or potential drug targets.

### Clinical Implications

The potential role of specific CNVs in treatment-resistance is also relevant to discussions surrounding the use of genetic testing in psychiatry. Genome wide screening technologies are routinely used to identify genomic etiologies in pediatric medical/neurodevelopmental settings and the benefits of testing in these populations are well established ^67, 68^. However, it remains an open question as to whether testing is similarly justified for other psychiatric conditions based on considerations of potential harms and benefits ^69^. As we have previously argued ^70, 71^, there is a shortage of the type of detailed clinical data needed to investigate the impact that large-effect CNVs have on health comorbidities and treatment outcomes in adult psychiatric patients. This limited knowledgebase has made it impossible to assess areas in which specific genetic etiologies may provide guidance to patients’ treatment management and understanding of individual health risks ^72^.

The 22q11.2 deletion syndrome (22q11DS) stands out among the group of SCZ CNVs, in that thorough investigations have been undertaken to characterize the associated phenotype in pediatric and adult carriers alike. As a result, clinical care guidelines have been established ^73^, and a diagnosis of 22q11DS even has implications for APD prescribing practices ^74^. These resources will become increasingly important in adult psychiatric practice as children diagnosed with 22q11DS (and other CNV syndromes) enter adulthood and seek out psychiatric care. We did not identify a significant increase of 22q11DS in our TRS cohort, which is consistent with the prevailing idea that psychosis associated with 22q11DS responds to APD treatment at rates similar to idiopathic schizophrenia ^75, 76^. Whether other SCZ CNVs (such as duplications of 15q11.2-q13.1 and 16p11.2) influence treatment-response, and through which biological pathways, should be the focus of future research efforts.

### Strengths and Limitations

There remains pervasive heterogeneity in the use of varying definitions of TRS in research. Since large, well-defined TRS cohorts are difficult to acquire, many studies base subject selection on surrogate measures of TRS, such as filled clozapine prescriptions or the use of polypharmacy, without any direct patient contact to confirm the TRS diagnoses or rule out instances of *pseudo-*TRS (i.e., medication noncompliance, substance abuse). Importantly, each subject in our TRS cohort resided in the protected environment of the PASH system for at least five years with continuous clinical care, medication management, adequate sustenance, and minimal access to substances of abuse.

Our approach to CNV detection (combining research-grade microarrays, exome sequencing, and clinical CLIA lab confirmation) also allowed us to maximize sensitivity while minimizing false positives. This combined approach may have captured CNVs that microarray alone would have missed, which must be considered when making comparisons of our findings to studies that used differing methods. For example, of the six 16p11.2 duplications identified in our TRS sample, only two were called from SNP-based microarray data, whereas all six were called from analysis of exome sequencing data and confirmed by a CLIA-certified lab. Larger studies will be necessary to confirm and further investigate the association of specific CNVs with TRS; however, this will likely necessitate significant collaborative efforts. Furthermore, there is a need for a greater depth of phenotypic data regarding adult psychiatric patients with disease-associated CNVs.

## Conclusions

In this carefully selected sample of patients with treatment resistant psychotic disorders, we identified an increased prevalence of CNVs at specific loci previously implicated in schizophrenia. These data support previous indications that disease-associated CNVs may be more common in treatment-resistant patients and offers two potential loci for further investigation: 15q11.2-q13.1 and 16p11.2. There is a need to carefully examine the global clinical outcomes, in psychiatry and other medical domains, across the lifespan of carriers of clinically relevant CNVs.

## Data Availability

All data produced in the present study are available upon reasonable request to the authors

## Data Availability

Data are available upon request from corresponding author R.C.J. and will also be available from national repositories with the NIMH study number of 150.

## Acknowledgements

The authors thank Dale Adair, MD, Chief Medical Officer, Office of Mental Health and Substance Abuse Services, Commonwealth of Pennsylvania, for his encouragement and support for this project. The authors also thank Takahiro Soda, MD, PhD, Department of Psychiatry, University of Florida, Gainesville Florida for reviewing the PASH cases in terms of inclusion/exclusion criteria. At the time of his assistance Dr. Soda held a position in the Department of Psychiatry, University of North Carolina (Chapel Hill). Marice Davis and Kelly Bingaman provided phlebotomy services with care and attention to detail for which we are grateful. We thank the NIMH and Rutgers University Cell and DNA Repository for support in sample collection and processing.

## References

1. Lally J, Gaughran F, Timms P, Curran SR. Treatment-resistant schizophrenia: current insights on the pharmacogenomics of antipsychotics. Pharmgenomics Pers Med 2016;9:117–129.

2. Howes OD, McCutcheon R, Agid O, et al. Treatment-Resistant Schizophrenia: Treatment Response and Resistance in Psychosis (TRRIP) Working Group Consensus Guidelines on Diagnosis and Terminology. Am J Psychiatry Mar 1 2017;174(3):216–229.

3. Iasevoli F, Giordano S, Balletta R, et al. Treatment resistant schizophrenia is associated with the worst community functioning among severely-ill highly-disabling psychiatric conditions and is the most relevant predictor of poorer achievements in functional milestones. Prog Neuropsychopharmacol Biol Psychiatry Feb 4 2016;65:34–48.

4. de Bartolomeis A, Balletta R, Giordano S, Buonaguro EF, Latte G, Iasevoli F. Differential cognitive performances between schizophrenic responders and non-responders to antipsychotics: correlation with course of the illness, psychopathology, attitude to the treatment and antipsychotics doses. Psychiatry Res Dec 15 2013;210(2):387–395.

5. Hor K, Taylor M. Suicide and schizophrenia: a systematic review of rates and risk factors. J Psychopharmacol Nov 2010;24(4 Suppl):81-90.

6. Kennedy JL, Altar CA, Taylor DL, Degtiar I, Hornberger JC. The social and economic burden of treatment-resistant schizophrenia: a systematic literature review. Int Clin Psychopharmacol Mar 2014;29(2):63–76.

7. Smart SE, Kepinska AP, Murray RM, MacCabe JH. Predictors of treatment resistant schizophrenia: a systematic review of prospective observational studies. Psychol Med Jan 2021;51(1):44–53.

8. Martin AK, Mowry B. Increased rare duplication burden genomewide in patients with treatment-resistant schizophrenia. Psychol Med Feb 2016;46(3):469–476.

9. Wimberley T, Gasse C, Meier SM, Agerbo E, MacCabe JH, Horsdal HT. Polygenic Risk Score for Schizophrenia and Treatment-Resistant Schizophrenia. Schizophr Bull Sep 1 2017;43(5):1064–1069.

10. Legge SE, Dennison CA, Pardinas AF, et al. Clinical indicators of treatment-resistant psychosis. Br J Psychiatry May 2020;216(5):259–266.

11. Kowalec K, Lu Y, Sariaslan A, et al. Increased schizophrenia family history burden and reduced premorbid IQ in treatment-resistant schizophrenia: a Swedish National Register and Genomic Study. Mol Psychiatry Nov 12 2019.

12. Pardinas AF, Smart SE, Willcocks IR, et al. Interaction Testing and Polygenic Risk Scoring to Estimate the Association of Common Genetic Variants With Treatment Resistance in Schizophrenia. JAMA Psychiatry Mar 1 2022;79(3):260–269.

13. Schizophrenia Working Group of the Psychiatric Genomics C. Biological insights from 108 schizophrenia-associated genetic loci. Nature Jul 24 2014;511(7510):421–427.

14. Szatkiewicz JP, O’Dushlaine C, Chen G, et al. Copy number variation in schizophrenia in Sweden. Mol Psychiatry Jul 2014;19(7):762–773.

15. Marshall CR, Howrigan DP, Merico D, et al. Contribution of copy number variants to schizophrenia from a genome-wide study of 41,321 subjects. Nat Genet Jan 2017;49(1):27–35.

16. Zoghbi AW, Dhindsa RS, Goldberg TE, et al. High-impact rare genetic variants in severe schizophrenia. Proc Natl Acad Sci U S A Dec 21 2021;118(51).

17. Singh T, Poterba T, Curtis D, et al. Rare coding variants in ten genes confer substantial risk for schizophrenia. Nature Apr 2022;604(7906):509–516.

18. Ruderfer DM, Charney AW, Readhead B, et al. Polygenic overlap between schizophrenia risk and antipsychotic response: a genomic medicine approach. Lancet Psychiatry Apr 2016;3(4):350–357.

19. Wang Q, Man Wu H, Yue W, et al. Effect of Damaging Rare Mutations in Synapse-Related Gene Sets on Response to Short-term Antipsychotic Medication in Chinese Patients With Schizophrenia: A Randomized Clinical Trial. JAMA Psychiatry Dec 1 2018;75(12):1261–1269.

20. Sriretnakumar V, Harripaul R, Vincent JB, Kennedy JL, So J. Enrichment of pathogenic variants in genes associated with inborn errors of metabolism in psychiatric populations. Am J Med Genet B Neuropsychiatr Genet Jan 2019;180(1):46–54.

21. Kirov G, Rees E, Walters JT, et al. The penetrance of copy number variations for schizophrenia and developmental delay. Biol Psychiatry Mar 1 2014;75(5):378–385.

22. Torres F, Barbosa M, Maciel P. Recurrent copy number variations as risk factors for neurodevelopmental disorders: critical overview and analysis of clinical implications. J Med Genet Feb 2016;53(2):73–90.

23. Rees E, Walters JT, Georgieva L, et al. Analysis of copy number variations at 15 schizophrenia-associated loci. Br J Psychiatry Feb 2014;204(2):108–114.

24. Kushima I, Aleksic B, Nakatochi M, et al. High-resolution copy number variation analysis of schizophrenia in Japan. Mol Psychiatry Mar 2017;22(3):430–440.

25. Ammons RB, Ammons CH. Quick Test.: Psychological Test Specialists; 1962.

26. Kay SR, Fiszbein A, Opler LA. The positive and negative syndrome scale (PANSS) for schizophrenia. Schizophr Bull 1987;13(2):261–276.

27. Harris PA, Taylor R, Minor BL, et al. The REDCap consortium: Building an international community of software platform partners. J Biomed Inform Jul 2019;95:103208.

28. Harris PA, Taylor R, Thielke R, Payne J, Gonzalez N, Conde JG. Research electronic data capture (REDCap)--a metadata-driven methodology and workflow process for providing translational research informatics support. J Biomed Inform Apr 2009;42(2):377–381.

29. Wang K, Li M, Hadley D, et al. PennCNV: an integrated hidden Markov model designed for high-resolution copy number variation detection in whole-genome SNP genotyping data. Genome Res Nov 2007;17(11):1665–1674.

30. Colella S, Yau C, Taylor JM, et al. QuantiSNP: an Objective Bayes Hidden-Markov Model to detect and accurately map copy number variation using SNP genotyping data. Nucleic Acids Res 2007;35(6):2013–2025.

31. Pinto D, Darvishi K, Shi X, et al. Comprehensive assessment of array-based platforms and calling algorithms for detection of copy number variants. Nat Biotechnol May 8 2011;29(6):512–520.

32. Zhang Z, Cheng H, Hong X, et al. EnsembleCNV: an ensemble machine learning algorithm to identify and genotype copy number variation using SNP array data. Nucleic Acids Res Apr 23 2019;47(7):e39.

33. Van der Auwera GA, Carneiro MO, Hartl C, et al. From FastQ data to high confidence variant calls: the Genome Analysis Toolkit best practices pipeline. Curr Protoc Bioinformatics 2013;43:11 10 11–11 10 33.

34. McLaren W, Gil L, Hunt SE, et al. The Ensembl Variant Effect Predictor. Genome Biol Jun 6 2016;17(1):122.

35. Fromer M, Moran JL, Chambert K, et al. Discovery and statistical genotyping of copy-number variation from whole-exome sequencing depth. Am J Hum Genet Oct 5 2012;91(4):597–607.

36. Pounraja VK, Jayakar G, Jensen M, Kelkar N, Girirajan S. A machine-learning approach for accurate detection of copy number variants from exome sequencing. Genome Res Jul 2019;29(7):1134–1143.

37. Rees E, Kendall K, Pardinas AF, et al. Analysis of Intellectual Disability Copy Number Variants for Association With Schizophrenia. JAMA Psychiatry Sep 1 2016;73(9):963–969.

38. Martin J, Tammimies K, Karlsson R, et al. Copy number variation and neuropsychiatric problems in females and males in the general population. Am J Med Genet B Neuropsychiatr Genet Sep 2019;180(6):341–350.

39. Kirov G, Grozeva D, Norton N, et al. Support for the involvement of large copy number variants in the pathogenesis of schizophrenia. Hum Mol Genet Apr 15 2009;18(8):1497–1503.

40. R: A language and environment for statistical computing [computer program]. Version: R Foundation for Statistical Computing; 2020.

41. epitools: Epidemiology Tools [computer program]. Version; 2020.

42. Budisteanu M, Papuc SM, Streata I, et al. The Phenotypic Spectrum of 15q13.3 Region Duplications: Report of 5 Patients. Genes (Basel) Jul 1 2021;12(7).

43. Gillentine MA, Berry LN, Goin-Kochel RP, et al. The Cognitive and Behavioral Phenotypes of Individuals with CHRNA7 Duplications. J Autism Dev Disord Mar 2017;47(3):549–562.

44. Pang H, Yu X, Kim YM, et al. Disorders Associated With Diverse, Recurrent Deletions and Duplications at 1q21.1. Front Genet 2020;11:577.

45. Vervoort L, Demaerel W, Rengifo LY, et al. Atypical chromosome 22q11.2 deletions are complex rearrangements and have different mechanistic origins. Hum Mol Genet Nov 15 2019;28(22):3724–3733.

46. Wada M, Noda Y, Iwata Y, et al. Dopaminergic dysfunction and excitatory/inhibitory imbalance in treatment-resistant schizophrenia and novel neuromodulatory treatment. Mol Psychiatry Apr 20 2022.

47. Mulle JG, Sullivan PF, Hjerling-Leffler J. Editorial overview: Rare CNV disorders and neuropsychiatric phenotypes: opportunities, challenges, solutions. Curr Opin Genet Dev Jun 2021;68:iii–ix.

48. Chan SKW, Chan HYV, Honer WG, et al. Predictors of Treatment-Resistant and Clozapine-Resistant Schizophrenia: A 12-Year Follow-up Study of First-Episode Schizophrenia-Spectrum Disorders. Schizophr Bull Mar 16 2021;47(2):485–494.

49. Ahn K, Gotay N, Andersen TM, et al. High rate of disease-related copy number variations in childhood onset schizophrenia. Mol Psychiatry May 2014;19(5):568–572.

50. Foley C, Heron EA, Harold D, et al. Identifying schizophrenia patients who carry pathogenic genetic copy number variants using standard clinical assessment: retrospective cohort study. Br J Psychiatry May 2020;216(5):275–279.

51. Modenato C, Kumar K, Moreau C, et al. Effects of eight neuropsychiatric copy number variants on human brain structure. Transl Psychiatry Jul 20 2021;11(1):399.

52. Seidlitz J, Nadig A, Liu S, et al. Transcriptomic and cellular decoding of regional brain vulnerability to neurogenetic disorders. Nat Commun Jul 3 2020;11(1):3358.

53. Urraca N, Cleary J, Brewer V, et al. The interstitial duplication 15q11.2-q13 syndrome includes autism, mild facial anomalies and a characteristic EEG signature. Autism Res Aug 2013;6(4):268–279.

54. Wolpert CM, Menold MM, Bass MP, et al. Three probands with autistic disorder and isodicentric chromosome 15. Am J Med Genet Jun 12 2000;96(3):365–372.

55. Shaaya EA, Pollack SF, Boronat S, Davis-Cooper S, Zella GC, Thibert RL. Gastrointestinal problems in 15q duplication syndrome. Eur J Med Genet Mar 2015;58(3):191–193.

56. Bernier R, Hudac CM, Chen Q, et al. Developmental trajectories for young children with 16p11.2 copy number variation. Am J Med Genet B Neuropsychiatr Genet Jun 2017;174(4):367–380.

57. Hudac CM, Bove J, Barber S, et al. Evaluating heterogeneity in ASD symptomatology, cognitive ability, and adaptive functioning among 16p11.2 CNV carriers. Autism Res Aug 2020;13(8):1300–1310.

58. Kim SH, Green-Snyder L, Lord C, et al. Language characterization in 16p11.2 deletion and duplication syndromes. Am J Med Genet B Neuropsychiatr Genet Sep 2020;183(6):380–391.

59. Steinman KJ, Spence SJ, Ramocki MB, et al. 16p11.2 deletion and duplication: Characterizing neurologic phenotypes in a large clinically ascertained cohort. Am J Med Genet A Nov 2016;170(11):2943–2955.

60. Filges I, Sparagana S, Sargent M, et al. Brain MRI abnormalities and spectrum of neurological and clinical findings in three patients with proximal 16p11.2 microduplication. Am J Med Genet A Aug 2014;164A(8):2003-2012.

61. Conant KD, Finucane B, Cleary N, et al. A survey of seizures and current treatments in 15q duplication syndrome. Epilepsia Mar 2014;55(3):396–402.

62. Bristow GC, Thomson DM, Openshaw RL, et al. 16p11 Duplication Disrupts Hippocampal-Orbitofrontal-Amygdala Connectivity, Revealing a Neural Circuit Endophenotype for Schizophrenia. Cell Rep Apr 21 2020;31(3):107536.

63. Rein B, Tan T, Yang F, et al. Reversal of synaptic and behavioral deficits in a 16p11.2 duplication mouse model via restoration of the GABA synapse regulator Npas4. Mol Psychiatry Jun 2021;26(6):1967–1979.

64. Willis A, Pratt JA, Morris BJ. BDNF and JNK Signaling Modulate Cortical Interneuron and Perineuronal Net Development: Implications for Schizophrenia-Linked 16p11.2 Duplication Syndrome. Schizophr Bull Apr 29 2021;47(3):812–826.

65. Frankle WG, Cho RY, Prasad KM, et al. In vivo measurement of GABA transmission in healthy subjects and schizophrenia patients. Am J Psychiatry Nov 1 2015;172(11):1148–1159.

66. Kaster TS, de Jesus D, Radhu N, et al. Clozapine potentiation of GABA mediated cortical inhibition in treatment resistant schizophrenia. Schizophr Res Jul 2015;165(2-3):157–162.

67. Miller DT, Adam MP, Aradhya S, et al. Consensus statement: chromosomal microarray is a first-tier clinical diagnostic test for individuals with developmental disabilities or congenital anomalies. Am J Hum Genet May 14 2010;86(5):749–764.

68. Srivastava S, Love-Nichols JA, Dies KA, et al. Meta-analysis and multidisciplinary consensus statement: exome sequencing is a first-tier clinical diagnostic test for individuals with neurodevelopmental disorders. Genet Med Nov 2019;21(11):2413–2421.

69. Morris E, O’Donovan M, Virani A, Austin J. An ethical analysis of divergent clinical approaches to the application of genetic testing for autism and schizophrenia. Hum Genet Aug 28 2021.

70. Farrell M, Lichtenstein M, Harner MK, et al. Treatment-resistant psychotic symptoms and the 15q11.2 BP1-BP2 (Burnside-Butler) deletion syndrome: case report and review of the literature. Transl Psychiatry Jan 28 2020;10(1):42.

71. Harner MK, Lichtenstein M, Farrell M, et al. Treatment-resistant psychotic symptoms and early-onset dementia: A case report of the 3q29 deletion syndrome. Schizophr Res Oct 2020;224:195–197.

72. Sullivan PF, Owen MJ. Increasing the Clinical Psychiatric Knowledge Base About Pathogenic Copy Number Variation. Am J Psychiatry Mar 1 2020;177(3):204–209.

73. Fung WL, Butcher NJ, Costain G, et al. Practical guidelines for managing adults with 22q11.2 deletion syndrome. Genet Med Aug 2015;17(8):599–609.

74. Mosheva M, Korotkin L, Gur RE, Weizman A, Gothelf D. Effectiveness and side effects of psychopharmacotherapy in individuals with 22q11.2 deletion syndrome with comorbid psychiatric disorders: a systematic review. Eur Child Adolesc Psychiatry Aug 2020;29(8):1035–1048.

75. Dori N, Green T, Weizman A, Gothelf D. The Effectiveness and Safety of Antipsychotic and Antidepressant Medications in Individuals with 22q11.2 Deletion Syndrome. J Child Adolesc Psychopharmacol Feb 2017;27(1):83–90.

76. Butcher NJ, Fung WL, Fitzpatrick L, et al. Response to clozapine in a clinically identifiable subtype of schizophrenia. Br J Psychiatry Jun 2015;206(6):484–491.

77. Goldstein JI, Jarskog LF, Hilliard C, et al. Clozapine-induced agranulocytosis is associated with rare HLA-DQB1 and HLA-B alleles. Nat Commun Sep 4 2014;5:4757.

78. O’Leary NA, Wright MW, Brister JR, et al. Reference sequence (RefSeq) database at NCBI: current status, taxonomic expansion, and functional annotation. Nucleic Acids Res Jan 4 2016;44(D1):D733–745.

79. Sjostedt E, Zhong W, Fagerberg L, et al. An atlas of the protein-coding genes in the human, pig, and mouse brain. Science Mar 6 2020;367(6482).

