## Supplemental Material for "Increased Prevalence of Rare Copy Number Variants in Treatment-Resistant Psychosis"

### Supplemental Methods

**1. Definition of Treatment Resistant Psychotic Symptoms**
For this study, "*Treatment Resistant Psychotic Symptoms"* (TRS) is defined as a lack of clinical improvement to at least three adequate trials of antipsychotic drugs (APDs) over at least five years of continuous hospitalization. Commonly defined as *"treatment resistant schizophrenia"*, here *"TRS"* indicates Treatment Resistant Symptoms of psychosis in schizophrenia as well as the related psychotic disorders (schizoaffective disorder, bipolar disorder with psychotic features, major depressive disorder with psychotic features, and psychosis NOS).

"*Lack of clinical improvement*" was determined from Global Assessment of Functioning (GAF) scores if available (consistent scores of ≤ 40 indicated an insufficient clinical response) [1]. If GAF scores were not available, the principal investigator (R.C.J.) discussed the case with the treating psychiatrist to confirm a lack of clinical improvement.

"*Continuous hospitalization*" was defined as uninterrupted inpatient treatment in a Pennsylvania State Hospital (PASH), and/or an affiliated Long Term Structured Residence (LTSR) facility. Many patients were transferred during their course of hospitalization (between PASHs, LTSRs, medical hospitals, and jail/forensic units). When transferred with uninterrupted treatment and supervision, these events were considered part of a continuous hospitalization. Any discharge into the community lasting more than 30 days was considered an interruption in continuous care, and subsequent re-admission would be considered the start of a new period of continuous hospitalization. In this way, risk of medication non-compliance and illicit drug use could be minimized.

An "*adequate trial"* was defined as exposure to an APD over a period of at least six weeks with daily doses equal to or greater than the defined daily dose (DDD) published by the World Health Organization (www.whocc.no). See **Supplemental Table S1** below for a complete list of APDs and their associated DDDs. Chlorpromazine equivalence is given as published in Woods, et al. [2], except where otherwise indicated.

**2. Determining Treatment Resistant Status**
Participants who signed an informed consent form were screened using the inclusion/exclusion criteria to determine TRS status. Often, criteria could be confirmed using hospital unit charts. In some cases, additional records were necessary for determining eligibility. For participants recruited from PASH, as well as those recruited from LTSRs who had previous hospitalizations in PASH, electronic pharmacy records were acquired which documented all medications administered from the beginning of the electronic record system in 1991 through the time of enrollment. These data included start/stop dates for each drug, as well as daily doses and mode of administration. For participants recruited from LTSRs without previous state hospital admissions,

or for participants who resided in the PASH before 1991, paper records were acquired whenever available. This included historical records from PASH hospitalizations, records from the LTSR where the participant was recruited, as well as other community or out of state hospitalizations. From paper records, start/stop dates and daily doses were extracted and used to confirm TRS status. Only drug trials during inpatient hospitalization (or stays in LTSRs) were considered in determining TRS status to ensure medication compliance.

**Supplemental Table S1: Defined Daily Dose and Chlorpromazine (CPZ) Equivalence**.

| Chemical Name | Defined Daily Dose (mg) | CPZ Equivalence(mg) |
|---|---:|---:|
| aripiprazole | 15 | 7.5 |
| asenapine** | 20 | 5 |
| chlorpromazine | 300 | 100 |
| clozapine | 300 | 50 |
| fluphenazine | 10 | 2 |
| haloperidol | 8 | 2 |
| Iloperidone** | 18 | 6 |
| loxapine | 100 | 10 |
| lurasidone** | 60 | 20 |
| mesoridazine | 200 | 50 |
| molindone | 50 | 10 |
| olanzapine | 10 | 5 |
| paliperidone** | 6 | 1.5 |
| perphenazine | 30 | 8 |
| pimozide* | 4 | 2 |
| prochlorperazine | 100 | 15 |
| quetiapine | 400 | 75 |
| risperidone | 5 | 2 |
| thioridazine | 300 | 100 |
| thiothixene | 30 | 4 |
| trifluoperazine | 20 | 2 |
| ziprasidone | 80 | 60 |
| haloperidol (depot) | 3.3 mg daily (92.4 mg q 4 weeks) | 30 mg q 4 weeks |
| fluphenazine (depot) | 1 mg daily (21 mg q 3 weeks) | 2.5 mg q 3 weeks |
| risperidone (depot) | 2.7 mg daily (37.8 mg q 2 weeks) | 25 mg q 2 weeks |

*https://cpnp.org/guideline/essentials/antipsychotic-dose-equivalents
**http://scottwilliamwoods.com/equivalencesupdate.php

Due to variations in record keeping between institutions, some patient records were unobtainable. These cases were reviewed carefully by the research team to determine final inclusion or exclusion. After discussions with the participant's treating physician, family member interviews, and review of all available records, the research team chose to include five participants in the final sample with only two *documented* adequate trials. These five participants each had long histories of psychotic illness with no indication of treatment response. All five cases had **three or more** APD trials, but only **two** were documented _with exact doses and durations_ in each case. Therefore, these patients would be considered TRS based on commonly accepted definitions for TRS [3] which require two adequate trials.

#### 3. CNV Calling Pipeline

Supplemental **Figure S1** visualizes the steps taken to identify CNVs within the sample. Preliminary chromosomal microarray (CMA) calls were filtered through Penn CNV to identify "likely pathogenic" CNVs which were then sent for independent verification in a clinical laboratory improvement amendment (CLIA)-certified lab. In two cases, including one with a mosaic copy loss of the Y chromosome (considered an incidental finding in this report) and a 16p13.11 duplication, subsequent analysis of CMA and whole exome sequencing (WES) data did not identify the calls (because of more stringent calling requirements and/or QC measures). However, both preliminary calls were confirmed by the CLIA lab, and thus are likely to represent real findings. This highlights the risk for type two error when using only one mode of CNV analysis. Likewise, CN-Learn identified six CNVs which were not called by the primary CMA pipeline. Again, those considered "likely-pathogenic" (n = 4; 16p11.2 dups) were sent for verification in an independent CLIA-certified clinical laboratory. Importantly, all CNVs identified with research-grade CMA and/or WES data which were sent for clinical confirmation (n=30) were also found by the CLIA laboratory, indicating a low risk for type one error in our methods.

**Supplemental Figure S1: CNV Analysis Work Flow.**

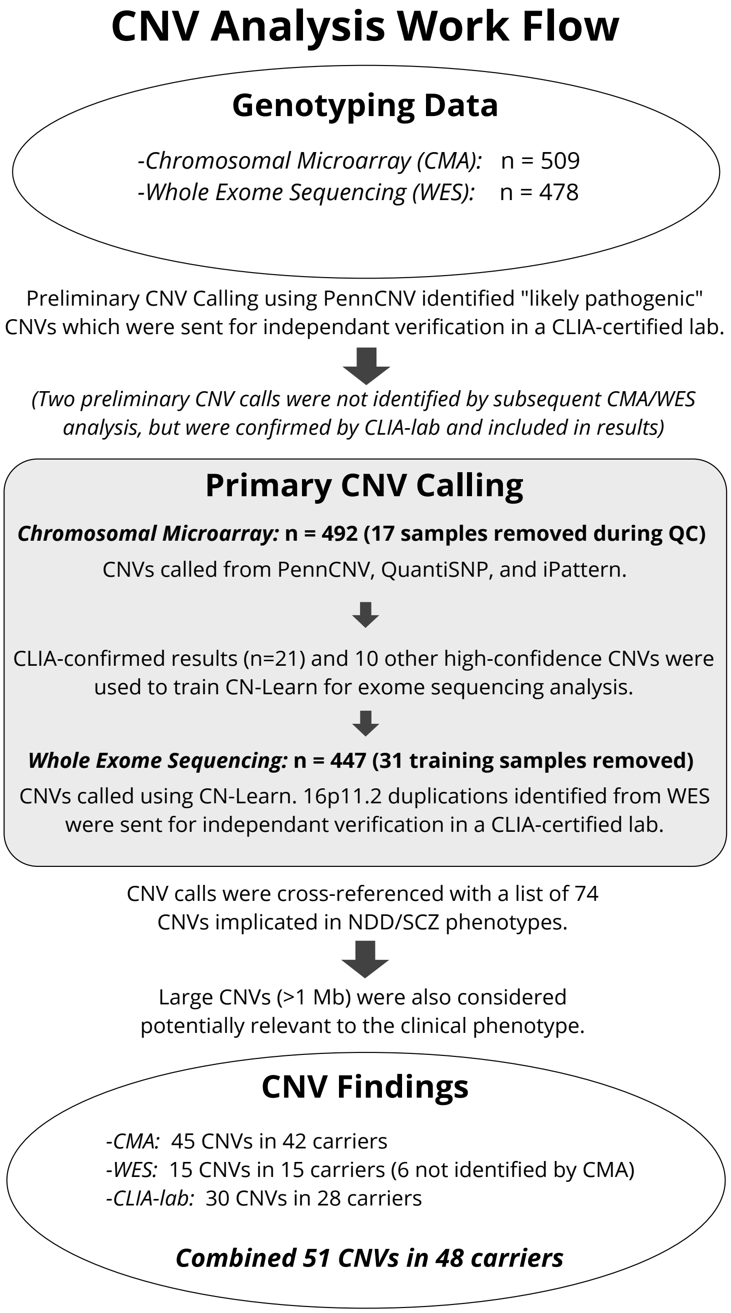

### 4. Grouping Copy Number Variants of Interest

CNVs most likely to have clinical significance were prioritized for investigation. To identify clinically relevant CNVs, we cross-referenced CNV calls from the TRS cohort with a curated list of 74 CNVs previously associated with neurodevelopmental or psychiatric disease (see **Supplemental Table S2**, below). This list of NDD variants was curated by Martin et al. [4]. Twelve schizophrenia-associated CNVs are also denoted, which were previously implicated by large case-control studies [5, 6].

CNVs were then categorized into one of three groups:

***1) Neurodevelopmental CNVs (NDD CNVs):*** Called CNVs overlapping known NDD CNV loci by more than 50% (see **supplemental table 2** for canonical breakpoint coordinates).

***2) Large CNVs not associated with neurodevelopmental phenotypes:*** CNVs greater than 1 MB (one million base pairs) are considered potentially relevant to pathology when identified in an individual with neurodevelopmental or psychiatric phenotypes. While case reports may describe individuals with CNVs in similar regions, no case-control studies have associated these CNVs with NDD or psychiatric phenotypes. **Supplemental Table S3** provides references for previously published reports of cases with CNVs of the same type, comparable size, and overlapping position as the cases identified in this report. References were identified using http://cnvxplorer.com/, and are not intended to be an exhaustive or all-encompassing search.

***3) Variants of Uncertain Significance in genomic areas of interest:*** This category encompasses two distinct subcategories of variants: 1) Called CNVs which partially overlap known NDD CNV risk loci, but with less than 50% overlap; 2) duplications at loci where deletions are known to be associated with NDD or psychiatric phenotypes, but where duplications have yet to be associated with increased disease risk.

### Supplemental Table S2: Known Neurodevelopmental CNV Loci from Previous Case-Control Studies.

| Locus/syndrome | Genomic Coordinates (hg19) | Size (bps) | CN type | SCZ-associated |
|---|---|---|---|---|
| 1p36 | chr1:telomere - 2,500,000 | 2,500,000 | deletion | |
| | | | duplication | |
| 1q21.1; TAR-syndrome region | chr1:145,394,955 - 145,807,817 | 412,862 | deletion | |
| | | | duplication | |
| **1q21.1** | **chr1:146,527,987 - 147,394,444** | **866,457** | **deletion** | **x** |
| | | | **duplication** | **x** |
| 1q24 (FMO and DNM3) | chr1:169,680,333 - 173,303,337 | 3,623,004 | deletion | |
| **2p16.3 (NRXN1)** | **chr2:50,145,643 - 51,259,674** | **1,114,031** | **deletion** | **x** |
| proximal 2p15-16.1 (PEX13 to AHSA2) | chr2:61,245,288 - 61,414,572 | 169,284 | duplication | |
| 2q11.2 | chr2:96,742,409 - 97,677,516 | 935,107 | deletion | |
| 2q13 | chr2:111,394,040 - 112,012,649 | 618,609 | deletion | |
| | | | duplication | |
| 2q33.1 (SATB2) | chr2:200,134,224 - 200,325,255 | 191,031 | deletion | |
| 2q37 (HDAC4) | chr2:239,716,679 - 243,199,373 | 3,482,694 | deletion | |
| 3p25.3 (JAGN1 to TATDN2) | chr3:9,932,271 - 10,322,902 | 390,631 | duplication | |
| 3p11.2 (CHMP2B to POU1F1) | chr3:87,267,612 - 87,531,631 | 264,019 | deletion | |
| 3q13 (GAP43) | chr3:115,332,334 - 115,504,038 | 171,704 | deletion | |
| 3q28-29 (FGF12) | chr3:191,859,728 - 192,126,012 | 266,284 | deletion | |
| **3q29** | **chr3:195,720,167 - 197,354,826** | **1,634,659** | **deletion** | **x** |
| 4p16.3; Wolf-Hirschorn region | chr4:1,552,030 - 2,091,303 | 539,273 | deletion | |
| | | | duplication | |
| 4q21 (BMP3) | chr4:81,945,477 - 81,985,327 | 39,850 | deletion | |
| 5q14 (MEF2C) | chr5:88,011,654 - 88,200,703 | 189,049 | deletion | |
| 5q35.3; Sotos syndrome | chr5:175,720,924 - 177,052,594 | 1,331,670 | deletion | |
| 7q11.23; William-Beuren syndrome region | chr7:72,744,915 - 74,142,892 | 1,397,977 | deletion | |
| | | | **duplication** | **x** |
| 7q11.23 | chr7:72,773,570 - 73,158,061 | 384,491 | deletion | |
| | | | duplication | |
| 7q11.23 | chr7:73,978,801 - 74,144,177 | 165,376 | deletion | |
| | | | duplication | |
| 7p36.3 (VIPR2, WDR60) | chr7:158,453,198 - 158,972,237 | 519,039 | deletion | suggestive risk |
| | | | duplication | |
| 8p23.1 | chr8:8,098,990 - 11,872,558 | 3,773,568 | deletion | |
| | | | duplication | |
| 8q22.2 (VPS13B) | chr8:100,025,494 - 100,889,808 | 864,314 | deletion | suggestive risk |
| 9p24.3 (DMRT1) | chr9:841,690 - 969,090 | 127,400 | deletion | suggestive risk |
| | | | duplication | suggestive risk |

### Supplemental Table S2 Continued.

| | | | | |
|---|---|---|---|---|
| 9p13 | chr9:32,648,800 - 38,808,255 | 6,159,455 | duplication | |
| 9q34 | chr9:138,460,697 - 141,036,426 | 2,575,729 | duplication | |
| 10q11.21q11.23 | chr10:49,390,199 - 51,058,796 | 1,668,597 | duplication | |
| 10q23 | chr10:82,045,472 - 88,931,651 | 6,886,179 | deletion | |
| 11p11.2; Potocki-Shaffer syndrome | chr11:43,940,000 - 46,020,000 | 2,080,000 | deletion | |
| 12p13 (SCNN1A to PIANP) | chr12:6,471,959 - 6,825,955 | 353,996 | duplication | |
| 15q11.2q13.1; Prader-Willi syndrome/ Angelman syndrome region | chr15:22,805,313 - 28,390,339 | 5,585,026 | deletion | |
| | | | **duplication** | **x** |
| **15q11.2** | **chr15:22,805,313 - 23,094,530** | **289,217** | **deletion** | **x** |
| 15q12 | chr15:26,971,834 - 27,548,820 | 576,986 | deletion | |
| **15q13.3 (CHRNA7)** | **chr15:31,080,645 - 32,462,776** | **1,382,131** | **deletion** | **x** |
| 15q13.3 | chr15:30943512-32515849 | 1,572,337 | duplication | |
| 15q24 | chr15:72,900,171 - 78,151,253 | 5,251,082 | deletion | |
| | | | duplication | |
| 15q25 | chr15:85,139,815 - 85,716,624 | 576,809 | deletion | |
| 16p13.11 | chr16:15,511,655 - 16,293,689 | 782,034 | deletion | |
| | | | **duplication** | **x** |
| 16p12.1 | chr16:21,950,135 - 22,431,889 | 481,754 | deletion | |
| **distal 16p11.2** | **chr16:28,823,196 - 29,046,783** | **223,587** | **deletion** | **x** |
| | | | duplication | |
| 16p11.2 | chr16:29,650,840 - 30,200,773 | 549,933 | deletion | |
| | | | **duplication** | **x** |
| 17p13.3 (YWHAE and PAFAH1B1) | chr17:1,247,834 - 2,588,909 | 1,341,075 | deletion | |
| | | | duplication | |
| 17p11.2; Smith-Magenis syndrome/ Potocki-Lupski syndrome region | chr17:16,812,771 - 20,211,017 | 3,398,246 | deletion | |
| | | | duplication | |
| 17q11.2 | chr17:29,107,491 - 30,265,075 | 1,157,584 | deletion | |
| | | | duplication | |
| 17q12 | chr17:34,815,904 - 36,217,432 | 1,401,528 | deletion | |
| | | | duplication | |
| 17q21.31; Koolen-de Vries syndrome | chr17:43,705,356 - 44,164,691 | 459,335 | deletion | |
| **22q11.2; DiGeorge/VCFS syndrome region** | **chr22:19,037,332 - 21,466,726** | **2,429,394** | **deletion** | **x** |
| | | | duplication | |
| distal 22q11.2 | chr22:21,920,127 - 23,653,646 | 1,733,519 | deletion | |
| | | | duplication | |
| 22q13.33; Phelan-McDermid syndrome region | chr22:51,113,070 - 51,171,640 | 58,570 | deletion | |
| | | | duplication | |

### Supplemental Table S3: Large (>1Mb) CNVs not implicated in neurodevelopmental phenotypes in case-control studies.

| CNV | Cases (N) | Genomic coordinates (size) | CNS genes [a] | Source | CLIA lab annotation [b] | Previously reported phenotypes (references) [c] |
|---|---|---|---|---|---|---|
| 2p12-p11.2 Del | 1 | chr2:75.99-84.39 (8.4 Mb) | CTNNA2 | CMA | - | CHD, DD, ID, DF, gait abnormalities; [7] |
| 4q28.3 Dup | 1 | chr4:131.71-133.77 (2.1 Mb) | - | CMA | - | Clonal B-cell lymphocytosis, essential thrombocythemia; [8] |
| 4q33 Dup | 1 | chr4:171.04-172.04 (1.0 Mb) | - | CMA | - | - |
| 5p14.3 Del | 1 | chr5:20.26-23.03 (2.8 Mb) | - | CMA | - | DD, LD, behavioral problems; [9] |
| 5q23.2 Del | 1 | chr5:125.14-126.33 (1.2 Mb) | ALDH7A1, LMNB1 | CMA, CLIA | Unclear Clinical Significance | Adult-onset leukoencephalopathy, cognitive impairment, pyridoxine-dependent epilepsy; [10, 11] |
| 6q22.31 Dup | 1 | chr6:117.99-120.65 (2.7 Mb) | CEP85L, NUS1 | CMA, WES | - | DD, cardiomyopathy; [12] |
| 7q21.13 Dup | 2 [d] | chr7:88.17-90.17 (2.0 Mb) | - | CMA | - | Cerebral palsy, CA; [13, 14] |
| 9q33.1 Dup | 1 | chr9:119.92-122.03 (2.1 Mb) | - | CMA | - | NDD (NOS), ASD; [15, 16] |
| 13q33.1-q34 Dup | 1 | chr13:104.45-115.11 (10.7 Mb) | LIG4, COL4A1, COL4A2, NAXD, GRK1, CHAMP1 | CMA, WES | - | CA, ID, DF, tall stature, psychosis, dystonia, breast cancer; [17, 18] |
| 17q25.3 Dup | 1 | chr17:76.41-78.94 (2.5 Mb) | CANT1, CCDC40, GAA, EIF4A3, SGSH, RNF213 | WES | - | CA, DF, neurological problems (infantile seizures, cerebral palsy), DD/ID, ADHD, anxiety; [8, 19-21] |
| 18p11.31-p11.23 Dup | 1 | chr18:6.84-8.05 (1.2 Mb) | LAMA1 | CMA, WES | - | Brain and ophthalmological malformations, DF, DD, ID, auditory impairment, short stature, GH deficiency; [22] |
| Total case count (% of TRS sample, *n* = 509) | 12 (2.4%) | | | | | |

Genomic coordinates aligned to hg19 (https://www.ncbi.nlm.nih.gov/grc); CNS, central nervous system; Mb, megabases; CHD, congenital heart disease; DD, developmental delay; ID, intellectual disability; DF, dysmorphic features; LD, learning disability; NDD, neurodevelopmental disorder; NOS, not otherwise specified; CA, congenital anomalies; ADHD, attention deficit-hyperactivity disorder; TRS, treatment resistant psychotic symptoms; GH deficiency, growth hormone deficiency. One case with a CNV >1Mb at the 16p centromere was not included here as this region has been documented as unlikely to be sensitive to copy number dosage changes (chr16:34,197,143-35,257,261; https://dosage.clinicalgenome.org/). Another case with a recurrent 17p12 deletion (pathogenic for Hereditary Neuropathy with Liability to Pressure Palsies) was considered an incidental finding and not reported here.

[a] Genes in CNV region implicated in CNS or developmentally related disease based on OMIM and ClinVar data.

[b] Interpretation of the clinical significance of CNVs provided by allele diagnostics based on review of available literature at the time of CNV call.

[c] Published reports of cases with CNVs of the same type, comparable size, and overlapping position as identified in this report, identified using http://cnvxplorer.com/, not intended to be an exhaustive or all-encompassing search.

[d] One carrier of the 7q21.13 duplications is also the carrier of the 2p12p11.2 deletion.

### References


1. American Psychiatric Association., American Psychiatric Association. Work Group to Revise DSM-III. *Diagnostic and statistical manual of mental disorders : DSM-III-R*. 3rd ed. Washington, DC: American Psychiatric Association; 1987.
2. Woods SW. Chlorpromazine equivalent doses for the newer atypical antipsychotics. *J Clin Psychiatry* Jun 2003;64(6):663-667.
3. Howes OD, McCutcheon R, Agid O, et al. Treatment-Resistant Schizophrenia: Treatment Response and Resistance in Psychosis (TRRIP) Working Group Consensus Guidelines on Diagnosis and Terminology. *Am J Psychiatry* Mar 1 2017;174(3):216-229.
4. Martin J, Tammimies K, Karlsson R, et al. Copy number variation and neuropsychiatric problems in females and males in the general population. *Am J Med Genet B Neuropsychiatr Genet* Sep 2019;180(6):341-350.
5. Rees E, Walters JT, Georgieva L, et al. Analysis of copy number variations at 15 schizophrenia-associated loci. *Br J Psychiatry* Feb 2014;204(2):108-114.
6. Marshall CR, Howrigan DP, Merico D, et al. Contribution of copy number variants to schizophrenia from a genome-wide study of 41,321 subjects. *Nat Genet* Jan 2017;49(1):27-35.
7. Silipigni R, Cattaneo E, Baccarin M, Fumagalli M, Bedeschi MF. Rare interstitial deletion of chromosome 2p11.2p12. Report of a new patient with developmental delay and unusual clinical features. *Eur J Med Genet* Jan 2016;59(1):39-42.
8. Kim H, Jang W, Shin S, et al. Two cases of concurrent development of essential thrombocythemia with chronic lymphocytic leukemia, one related to clonal B-cell lymphocytosis, tested by array comparative genomic hybridization. *Int J Hematol* Jun 2015;101(6):612-619.
9. Barber JC, Huang S, Bateman MS, Collins AL. Transmitted deletions of medial 5p and learning difficulties; does the cadherin cluster only become penetrant when flanking genes are deleted? *Am J Med Genet A* Nov 2011;155A(11):2807-2815.
10. Nmezi B, Giorgio E, Raininko R, et al. Genomic deletions upstream of lamin B1 lead to atypical autosomal dominant leukodystrophy. *Neurol Genet* Feb 2019;5(1):e305.
11. Mefford HC, Zemel M, Geraghty E, et al. Intragenic deletions of ALDH7A1 in pyridoxine-dependent epilepsy caused by Alu-Alu recombination. *Neurology* Sep 1 2015;85(9):756-762.
12. Lee TM, Addonizio LJ, Chung WK. Dilated cardiomyopathy due to a phospholamban duplication. *Cardiol Young* Oct 2014;24(5):953-954.
13. McMichael G, Girirajan S, Moreno-De-Luca A, et al. Rare copy number variation in cerebral palsy. *Eur J Hum Genet* Jan 2014;22(1):40-45.
14. Molck MC, Simioni M, Paiva Vieira T, et al. Genomic imbalances in syndromic congenital heart disease. *J Pediatr (Rio J)* Sep - Oct 2017;93(5):497-507.
15. Lionel AC, Tammimies K, Vaags AK, et al. Disruption of the ASTN2/TRIM32 locus at 9q33.1 is a risk factor in males for autism spectrum disorders, ADHD and other neurodevelopmental phenotypes. *Hum Mol Genet* May 15 2014;23(10):2752-2768.
16. Chang YS, Lin CY, Huang HY, Chang JG, Kuo HT. Chromosomal microarray and whole-exome sequence analysis in Taiwanese patients with autism spectrum disorder. *Mol Genet Genomic Med* Dec 2019;7(12):e996.
17. Mak CCY, Chow PC, Liu APY, et al. De novo large rare copy-number variations contribute to conotruncal heart disease in Chinese patients. *NPJ Genom Med* 2016;1:16033.
18. Moscovich M, LeDoux MS, Xiao J, et al. Dystonia, facial dysmorphism, intellectual disability and breast cancer associated with a chromosome 13q34 duplication and overexpression of TFDP1: case report. *BMC Med Genet* Jul 13 2013;14:70.
19. Wang Q, Li Q, Xu Q, Liu Y, Yuan H. [Identification of a 17q25.3 duplication in a Chinese patient with global developmental delay and multiple congenital anomalies]. *Zhonghua Yi Xue Yi Chuan Xue Za Zhi* Jan 10 2020;37(1):52-56.

**20.** Probst FJ, James RA, Burrage LC, et al. De novo deletions and duplications of 17q25.3 cause susceptibility to cardiovascular malformations. *Orphanet J Rare Dis* Jun 14 2015;10:75.
**21.** Lukusa T, Fryns JP. Pure de novo 17q25.3 micro duplication characterized by micro array CGH in a dysmorphic infant with growth retardation, developmental delay and distal arthrogryposis. *Genet Couns* 2010;21(1):25-34.
**22.** Giordano M, Muratore V, Babu D, Meazza C, Bozzola M. A 18p11.23-p11.31 microduplication in a boy with psychomotor delay, cerebellar vermis hypoplasia, chorioretinal coloboma, deafness and GH deficiency. *Mol Cytogenet* 2016;9:89.
